# Intestinal fungal dynamics and linkage to hematopoietic cell transplantation outcomes

**DOI:** 10.1101/2021.07.20.21260859

**Authors:** Thierry Rolling, Bing Zhai, Mergim Gjonbalaj, Nicholas Tosini, Keiko Yasuma-Mitobe, Emily Fontana, Luigi A Amoretti, Roberta J Wright, Doris M Ponce, Miguel A Perales, Joao B Xavier, Marcel R M van den Brink, Kate A Markey, Jonathan U Peled, Ying Taur, Tobias M Hohl

## Abstract

Allogeneic hematopoietic cell transplantation (allo-HCT) induces profound shifts in the intestinal bacterial microbiota. The dynamics of intestinal fungi and their impact on clinical outcomes have not yet been integrated into a model of microbiota function during allo-HCT. Here, we combined parallel high-throughput fungal ITS1 amplicon sequencing, bacterial 16S amplicon sequencing, and fungal cultures to reveal striking trans-kingdom dynamics and their association with patient outcomes. We saw that the overall density and the biodiversity of intestinal fungi were stable during allo-HCT, but the species composition changed drastically from day to day. We identified a subset of patients with fungal dysbiosis characterized by culture positivity, stable expansion of *Candida parapsilosis* complex species, and distinct trans-kingdom microbiota profiles. These patients had worse overall survival and higher transplant-related mortality independent of candidemia. Our data expand the clinical significance of the mycobiota and suggest that targeting fungal dysbiosis may help to improve long-term patient survival.

---

Allogeneic hematopoietic cell transplantation (allo-HCT) is a curative treatment for certain malignant and non-malignant hematological disorders. Patients that undergo allo-HCT receive cytotoxic conditioning chemotherapy prior to infusion of donor immune cells. To prevent opportunistic infections during this period of intense immunosuppression, patients typically receive routine antibacterial and antifungal prophylaxis ^1-6^. The impact of allo-HCT on the bacterial microbiota has been characterized in prior studies: intestinal bacterial burden and α-diversity start to decline in the week prior to donor hematopoietic cell infusion, concurrent with the initiation of conditioning chemotherapy and antimicrobial therapy ^7-10^. Damage to the intestinal microbiota primarily affects anaerobic bacterial taxa that mediate colonization resistance against pathobionts, e.g., *Enterococcus* and *Enterobacteriaceae* ^10,11^. Low bacterial diversity and enterococcal domination have been associated with lower overall survival, higher transplant-related mortality, and higher graft-versus-host-related mortality in allo-HCT patients ^7,12^. Domination by bacterial pathobionts in the gut precedes bacterial translocation and the development of bloodstream infections ^10,13,14^.

In a recent case-control study of 15 patients using culture-dependent and -independent analyses of paired fecal samples and bloodstream fungal isolates, we reported that intestinal expansion of *Candida* species preceded bloodstream invasion, linking a perturbation in the fungal mycobiota to invasive fungal disease ^15^. In addition, exposure to antifungal prophylaxis and the presence of *Candida* colonization by culture-based surveys has been linked to acute intestinal graft versus host disease ^16,17^. In studies of inflammatory bowel disease and intestinal wound healing, a number of fungal species, including *Malassezia, Candida albicans*, and *Debaryomyces*, have been associated with colitis exacerbations, responsiveness to fecal microbiota transplantation, or defective wound healing ^18-20^, though it is unknown whether shifts in intestinal fungi predict long-term survival in inflammatory bowel disease or HCT.

Here, we characterize the composition and dynamic changes of intestinal fungal communities by both longitudinal amplicon-based and culture-dependent analyses in fecal samples collected from 156 allo-HCT patients. Allo-HCT was associated with a substantial increase in viable fungi that were recovered in fecal samples, in conjunction with an expansion of *Candida parapsilosis* complex species in a subset of patient. These patients had distinct trans-kingdom microbiota profiles. Patients with intestinal *C. parapsilosis* complex species expansion had a reduced overall survival that was explained by increased transplant-related mortality.

## Results

### Study characteristics

To analyze intestinal fungi during allo-HCT, we performed a cohort study that included 156 patients with 1279 prospectively collected fecal samples (median: 10 per patient). Collection started 10 days prior to and continued until 30 days after allo-HCT. These subjects were randomly drawn from a large-scale, prospective stool biobanking effort at our institution, which has been described previously ^7,10^. Patients underwent transplantation between January 1^st^, 2017, and December 31^st^, 2018, and had a median clinical follow-up of 32 months for this study (see cohort characteristics in Table 1). The most common indication for transplantation was acute leukemia and most patients received unmodified grafts. In this cohort, the median hematopoietic-cell transplantation comorbidity index (HCT-CI) risk score ^21^ was 2 (interquartile range: 1 to 4), corresponding to an intermediate mortality risk. The patients received antifungal prophylaxis with intravenous micafungin (150 mg/day), an echinocandin drug starting 7 days prior to HCT for a median of 17 days (IQR 15 to 22 days), prior to advancing to azole-based fungal prophylaxis, consistent with current institutional practices.

**Table 1:** Patient Characteristics.

| Patient Characteristics | N = 156 |
| --- | --- |
| Age at HCT (years, median [IQR]) | 60 (50-65) |
| Female Sex (n, %) | 63 (40%) |
| Primary Disease (n, %) |  |
| - Acute leukemia | - 80 (51%) |
| - Myelodysplastic syndrome | - 28 (18%) |
| - Lymphoma | - 25 (16%) |
| - Myeloproliferative disorder | - 10 (6%) |
| - Multiple myeloma | - 8 (5%) |
| - Other | - 5 (3%) |
| Graft Type (n, %) |  |
| - Unmodified PBSC | - 62 (40%) |
| - T-cell depleted PBSC <sup>a</sup> | - 47 (30%) |
| - Unmodified bone marrow | - 31 (20%) |
| - Umbilical cord blood | - 16 (10%) |
| HCT-CI (median, [IQR]) | 2 (1-4) |
| Follow-up (months, median [IQR]) | 32 (15-38) |

<sup>a</sup> one patient received a graft that was selectively depleted of alpha/beta-T-cells.

### The fungal mycobiota and bacterial microbiota exhibit distinct dynamics during allo-HCT

To assess the temporal dynamics of the intestinal mycobiota, we quantified fungal burden and diversity in longitudinally obtained samples. Fungal density was quantified by qPCR with the fungal 18S ribosomal DNA as a target ^22^. While the fecal fungal DNA content exhibited a large variation at all time points examined, there was no overall time-dependent change and no difference in fungal DNA abundance pre- and post-transplantation (Fig. 1a, Extended data Fig. 1a). Similarly, the fungal α-diversity at the species-level varied only marginally over the time course of allo-HCT (Fig. 1b, Extended data Fig. 1b). These results contrasted with a significant loss in bacterial density (Fig. 1c, Extended data Fig. 1c) and diversity (Fig. 1d, Extended data Fig. 1d) around the time of HCT.

**Figure 1:**
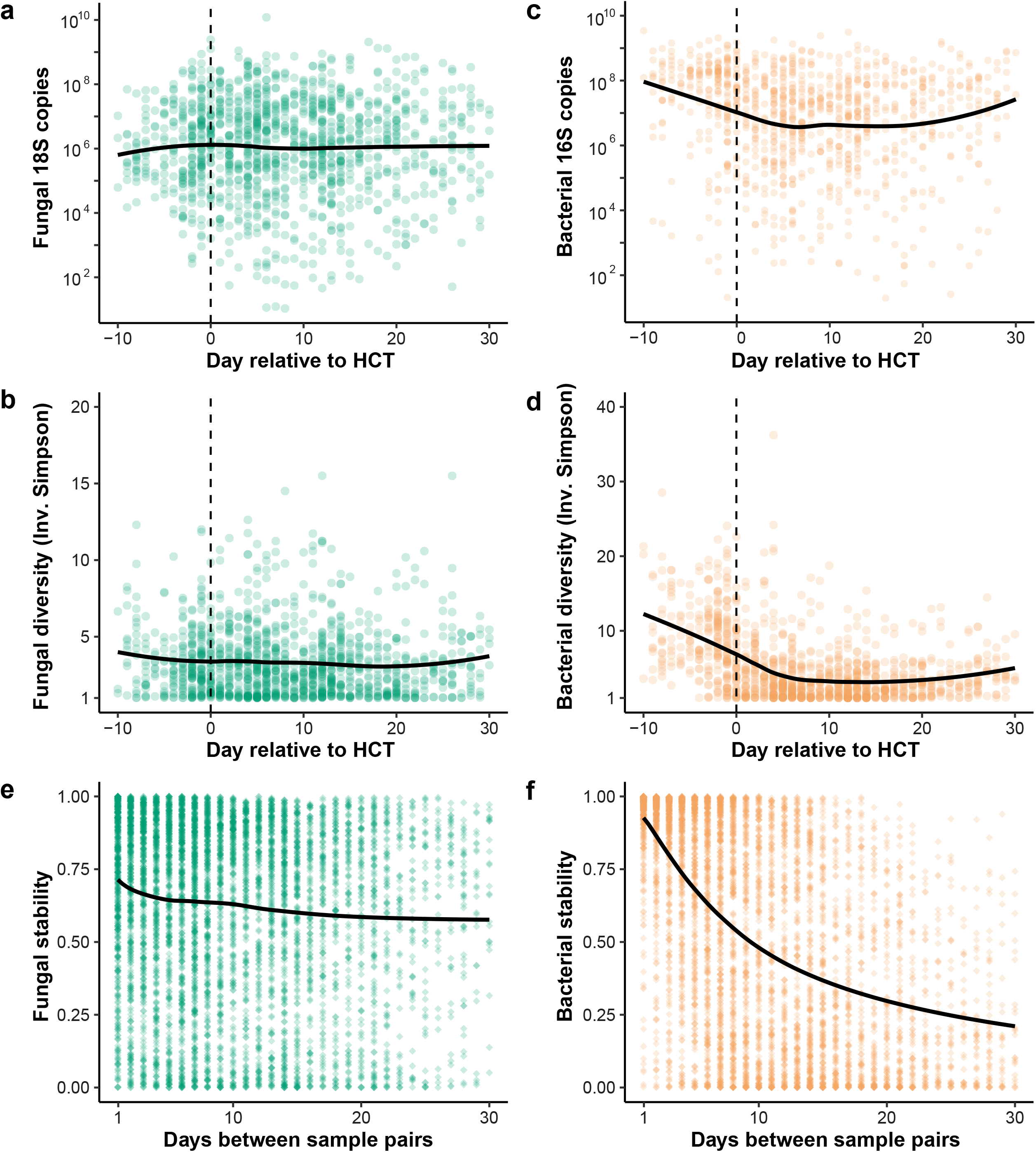
Dynamics of the fungal and bacterial microbiota during allo-HCT. (a) Fungal density measured by 18S qPCR (n = 1224 samples) and (b) fungal diversity at species level measured by the Inverse Simpson Index (n = 1279 samples) over time. (c) Bacterial density measured by 16S qPCR (n = 953 samples) and (d) bacterial diversity at species level measured by the Inverse Simpson Index (n = 1225 samples) over time. (a-d) The dashed line represents day 0, the day of donor cell infusion. Each dot represents a single sample. (e) Fungal (n = 5916 sample pairs) and (f) bacterial (n = 5171 sample pairs) stability during allo-HCT. The Chao-Jaccard index at species level. was used to calculate similarity between two samples from one individual. A value of 1 (1-Chao Jaccard) corresponds to two samples with identical composition, while a value of corresponds to two samples not sharing any species. (e-f) Each dot represents a sample pair from a single patient. Statistics: The solid black line corresponds to the smoothed average using Loess smoothing. See also Extended data Fig. 1.

The distinct temporal dynamics of the fungal and bacterial microbiota included a lower day-to-day stability, as measured by the Chao-Jaccard similarity index ^23^ of the fungal mycobiota composition compared to that of the bacterial microbiota (Fig. 1e and 1f). In the ensuing analysis, a value of 1 corresponds to an identical composition of 2 samples, while a value of 0 corresponds to no taxa being present in both samples. For sample pairs collected one day apart, the fungal mycobiota had a median stability (1-Chao-Jaccard) of 0.79, and the bacterial microbiota a median stability of 0.98. For samples collected 30 days apart, the fungal mycobiota had a median similarity of 0.73, and the bacterial microbiota had a median similarity of 0.19, consistent with prior reports of a decline in the similarity of the bacterial microbiota over time, best fitting to a power law ^24^. Thus, fecal samples collected on consecutive days differed substantially in their mycobiota composition and were not more similar to each other than two samples from the same patient collected 30 days apart. In contrast, two samples collected on consecutive days had a much more similar bacterial composition than two samples from the same patient collected 30 days apart.

### Fungal culture-positivity and expansion of single taxa define intestinal fungal dysbiosis

Dysbiosis of the gut mycobiota is not necessarily reflected in summary measures such as density and diversity. Thus, we further characterized the mycobiota composition by (1) analyzing culturable fungi recovered from fecal samples on Sabouraud agar supplemented with vancomycin and gentamicin, and (2) performing taxonomic profiling at the species level via analysis of internal transcribed spacer 1 (ITS1) amplicons that were derived from a segment of spacer DNA located between the fungal 18S and 5.8S rRNA genes ^25^. Culturable fungi were recovered from 195 out of 1279 samples (15%). The proportion of fecal samples from which fungi could be cultured was substantially higher in the post-transplant period (day 0 to +30) compared to the pre-transplant period (day -10 to -1; Fig. 2a). *C. parapsilosis* complex species (*C. parapsilosis, C. orthopsilosis, and C. metapsilosis)* were the most commonly cultured fungi, followed by *S. cerevisiae*, and other *Candida* species (Fig. 2b). The presence of viable fungi was associated with an increase in intestinal fungal biomass (Fig. 2c), and with a reduction in fungal α-diversity (Fig. 2d).

**Figure 2:**
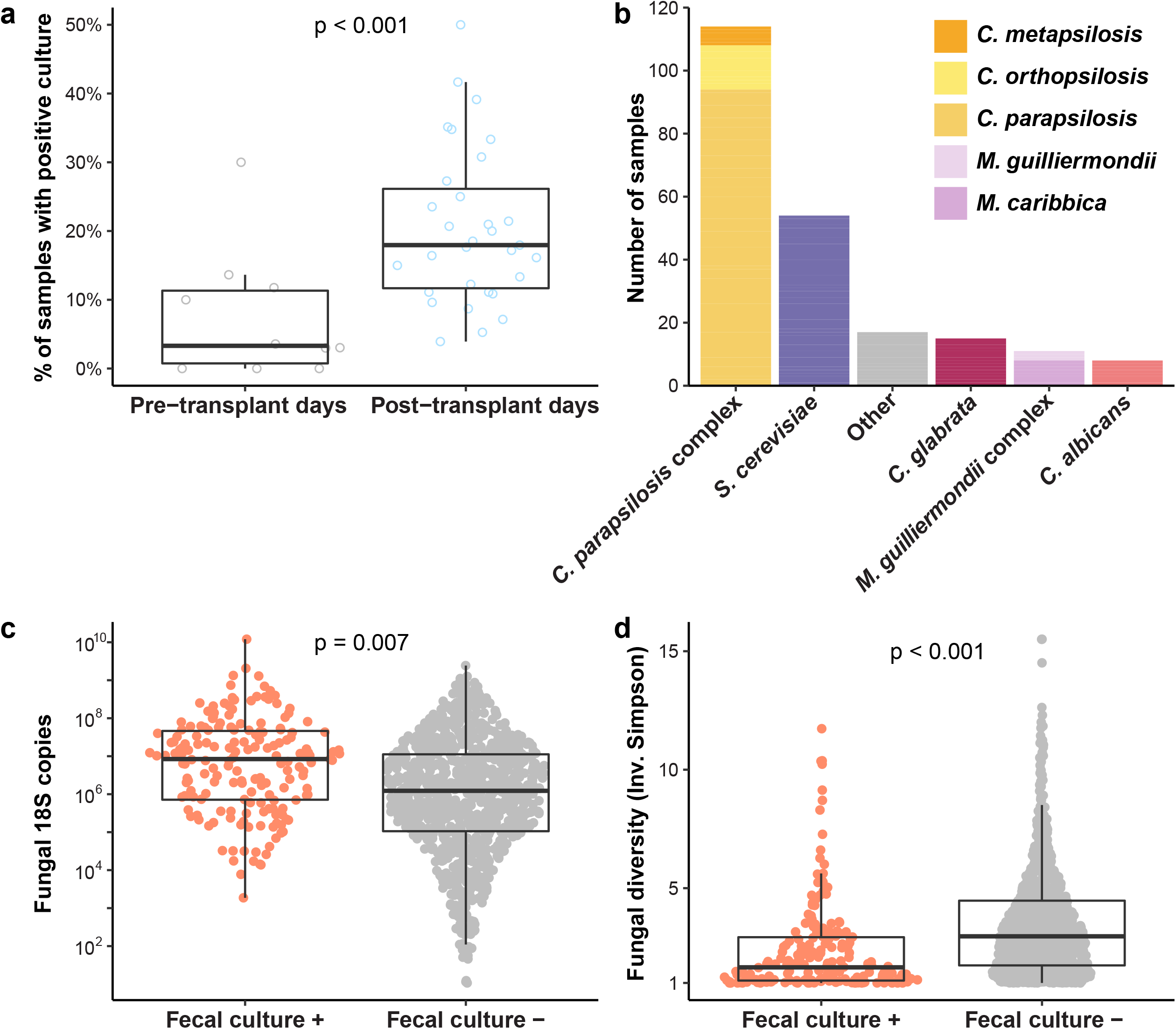
Distinct characteristics of fungal culture-positive and -negative fecal samples. (a) Percentage of fecal samples with a positive fungal culture per day. Each circle represents the proportion for a single day. The pre-transplant period is defined by days -10 to -1, the post-transplant period by days 0 to 30. P-value by Wilcoxon rank sum test. (b) Number of samples from which the specified fungal species could be cultured. The column labeled “other” groups fungal species found in five samples or less. Viable fungi were cultured from 195 samples and include 219 species. (c) Fungal density by 18S copies (n = 179 fecal culture positive samples and n= 1045 fecal culture negative samples) and (d) diversity by Inverse Simpson (n = 194 fecal culture positive samples and n=1085 fecal culture negative samples) in fecal samples with positive and negative fungal cultures. (c-d): P-values issued from generalized estimating equations with patient ID as cluster variable are shown.

To assess the relationship between culture-dependent and independent methods of sample analysis in our dataset, we compared the culture results to results obtained by ITS1 amplicon sequencing. In 177 of the 194 (91%) culture-positive samples, the cultured fungus could be detected in ITS1 sequences (Extended data Fig 2a). Figures 3A-3D show both the longitudinal culture results and the ITS1 amplicon composition of fecal samples from representative patients during allo-HCT. Data from representative subjects in Fig. 3a and 3b suggest that culture positivity and the relative expansion of corresponding taxa in the ITS1 sequences coincide temporally. To explore the association between culture-positivity and sequence expansion by ITS1 amplicon sequencing, we defined fungal domination by the presence of a single species at 90% relative abundance or higher. We found that if a sample was dominated by a fungal species on one day, the mycobiota composition of the next day was more similar than a pair of samples collected on consecutive days in which the first sample was not dominated by a single taxon (Extended data Fig 2b). This similarity in β-diversity was highest for initial samples dominated by *C. parapsilosis* complex, followed by samples dominated by *C. albicans* (Extended data Fig. 2c). This is consistent with our finding that domination by *C. parapsilosis* complex and subsequent culture-positivity could persist over several days (Fig 3a and b).

**Figure 3:**
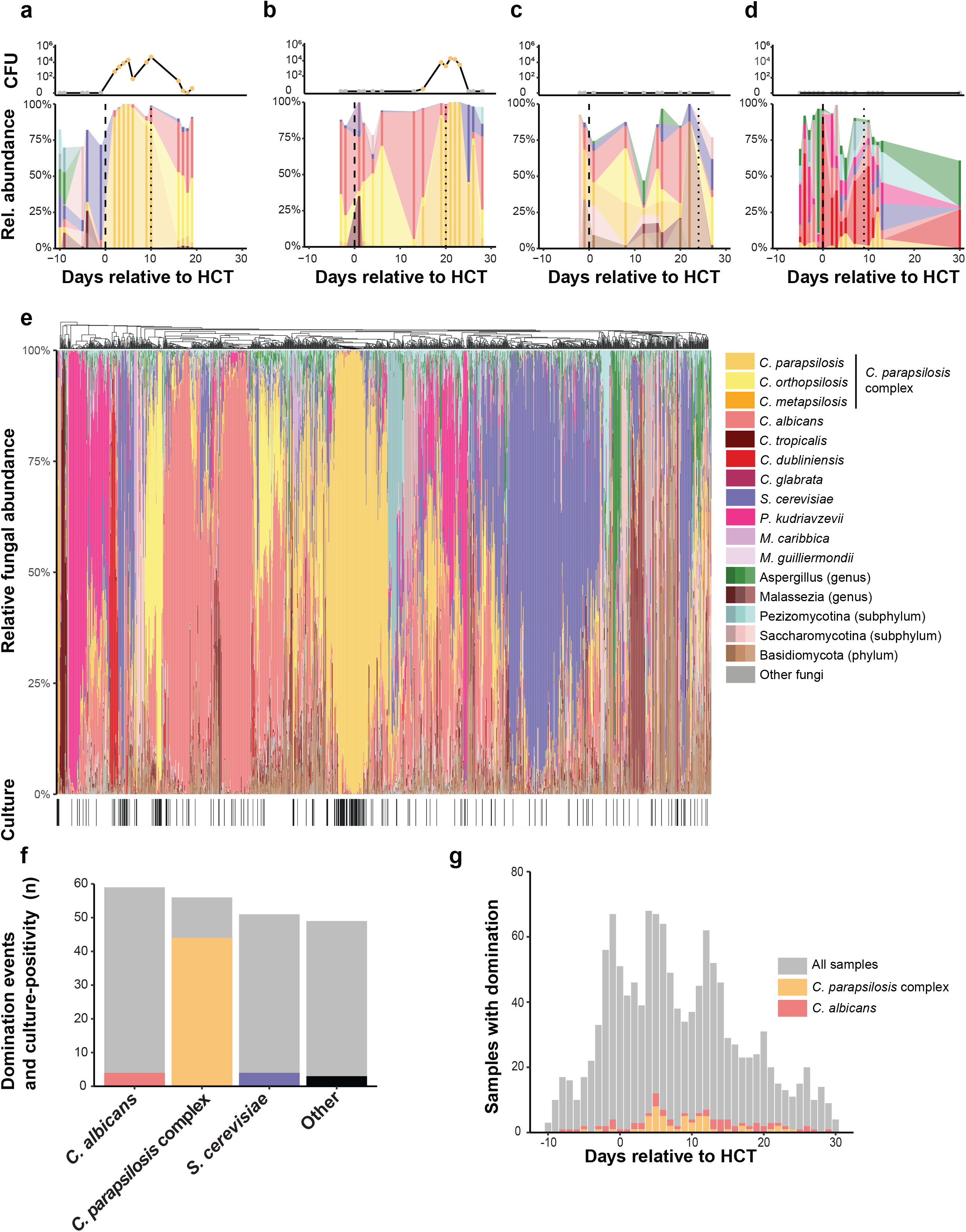
Fungal composition in fecal samples during allo-HCT. (a-d) Representative plots for individual patients (a, b) with viable fungi and (c, d) without viable fungi in fecal samples. The top rows depict the number of colony-forming units of fungi cultured from individual samples, color-coded by the cultured fungal species. The bottom rows depict the taxonomic composition of each sample. Fungal species present in a single sample or present below a peak abundance of 10% have been blanked in the graphs. (e) Hierarchical clustering at genus level of all samples and culture-positivity. A black line in the lower ribbon represents a positive culture from the respective sample (n = 1279 samples). (f) Domination events and relation to culture-positivity for major taxa. The grey bars represent the number of samples with domination (≥90% relative abundance) of the specific taxon. The colored bars represent the number of samples with domination and a positive culture by the specific taxon. (g) Histogram of the temporal relation of domination by *C. parapsilosis* complex and *C. albicans*. See also Extended data Fig. 2

Overall, 215 (17%) samples that belonged to 96 patients (62% of cohort) showed fungal domination with the 90% threshold in relative abundance (Fig 3e). *C. albicans* (59, 5%), *C. parapsilosis* complex species samples (56, 5%), *S. cerevisiae* (51, 4%), and *Pichia kudriavzevii (C. krusei)* (24, 2%) were the most common taxa that dominated fecal samples. Collectively, all other fungal taxa dominated less than 10 samples. Comparison of patients with culture-positive and culture-negative fecal samples revealed striking differences (Fig. 3e and 3f). Samples with *C. parapsilosis* complex species domination were culture positive in 43/56 (77%) cases. In contrast, culture positivity was rare for samples with *C. albicans* 10/59 (17%), *S. cerevisiae* 10/51 (20%), and *P. kudriavzevii* 3/24 (13%) domination (Fig. 3f). Domination events with *C. parapsilosis* complex species clustered in the immediate post-transplant period. In contrast, samples dominated by *C. albicans* were more evenly distributed across the study period (Fig. 3g). In sum, *C. parapsilosis* complex species expanded in the intestine in a subset of patients after HCT, and this domination state was frequently associated with the recovery of culturable fungi from fecal samples.

### Cross-kingdom interactions between intestinal fungi and bacteria in allo-HCT patients

To examine microbiota-intrinsic factors that led to fungal domination, we assessed intestinal trans-kingdom interactions between bacteria and fungi. Samples with *C. parapsilosis* complex domination had a substantially lower bacterial density than samples without any fungal domination. In contrast, *C. albicans* domination states were not associated with a lower bacterial biomass (Fig. 4a). Bacterial α-diversity did not discriminate between different fungal domination states (Fig. 4b). To assess whether the lack of association between bacterial diversity and fungal composition might be due to a different composition of the bacterial microbiota, we first assessed co-domination patterns between bacteria and fungi (Fig 4c). Most fungal domination states were not associated with concurrent bacterial domination states – commonly defined as presence of a species at 30% relative abundance or higher ^7,10,13^. Intriguingly, co-domination between *C. parapsilosis* complex and *Enterococcus* was extremely rare, although *Enterococcus* domination is one of the most common bacterial domination states in allo-HCT patients this cohort ^7,10-12^. In addition, co-domination events of *Staphylococcus* and *Streptococcus species* were more commonly observed with *C. parapsilosis* complex than with *C. albicans* or *S. cerevisiae*. To explore these observations further, we correlated changes in the bacterial and fungal mycobiota. Variations in the relative abundance of *C. parapsilosis* complex, *C. albicans*, and *S. cerevisiae* correlated with distinct bacterial signatures (Fig. 4c). Increases in *C. parapsilosis* complex abundance correlated with an increase in taxa belonging to the family Staphylococcaceae, and negatively correlated with Enterococcaceae (Fig. 4d), as expected from the co-domination analysis (Fig. 4c). The relative abundance of *C. parapsilosis* complex species correlated positively with Aerococcaceae, Flavobacteriaceae, Mycobacteriaceae, Enterobacteriaceae, Mycoplasmataceae, and Verrucomicrobiaceae. Importantly, *C. albicans* and *S. cerevisiae* were associated with distinct bacterial family correlation profiles compared to *C. parapsilosis* complex species (Fig 4d).

**Figure 4:**
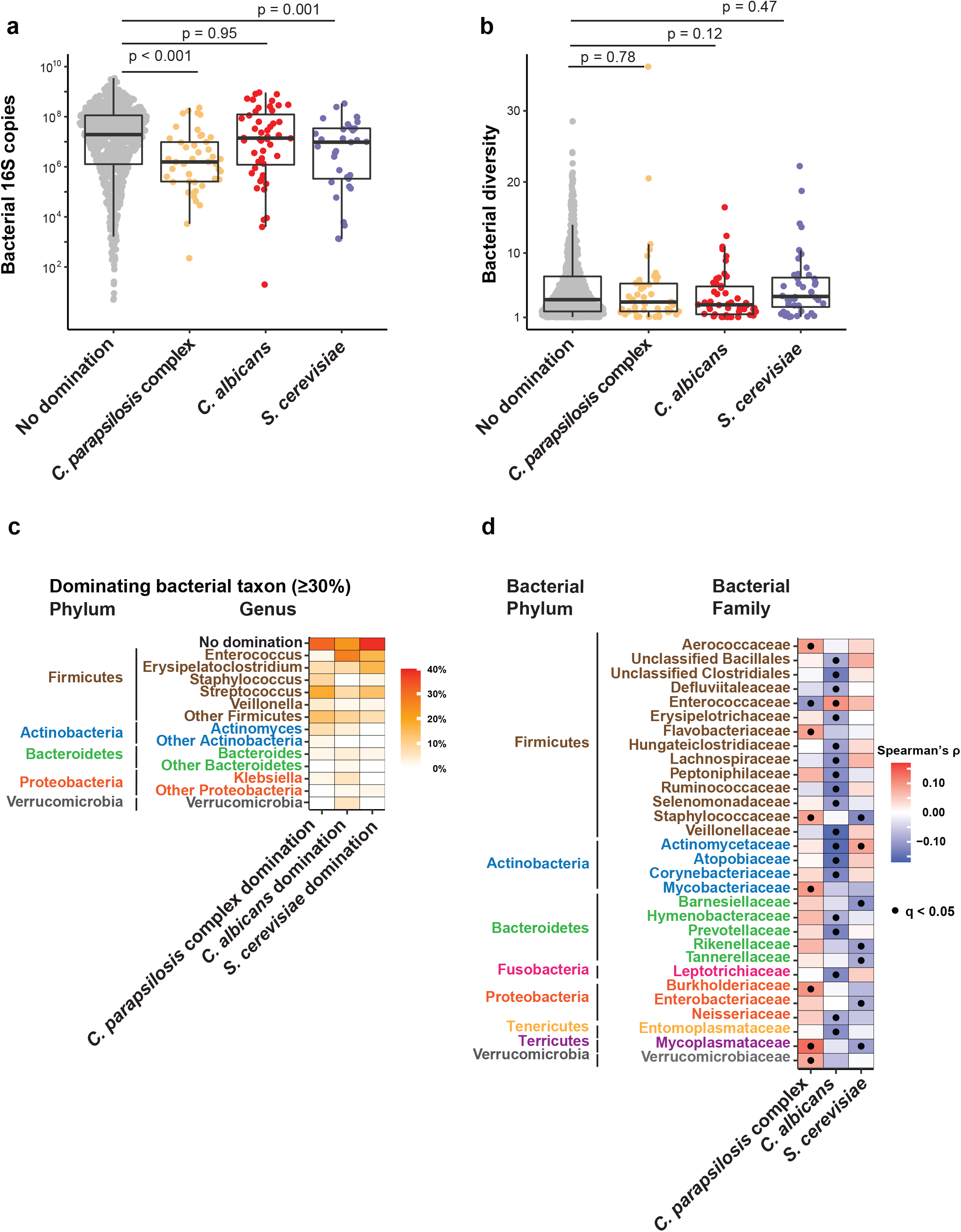
Trans-kingdom bacterial and fungal interactions during allo-HCT. (a) Comparison of bacterial density (n = 921 samples: 795 with no domination, 45 with *C. parapsilosis* complex domination, 48 with *C. albicans* domination, and 33 with *S. cerevisiae* domination) and (b) diversity (n = 1230 samples: 1064 with no domination, 56 with *C. parapsilosis* complex domination, 59 with *C. albicans* domination, and 51 with *S. cerevisiae* domination) in fecal samples with different dominating fungal taxa (≥ 90% relative abundance). Between group differences were evaluated by generalized estimating equations with patient ID as cluster variable. (c) Co-domination matrix between bacterial taxa (defined as relative abundance of a bacterial species ≥ 30%) and fungal domination (n = 140). (d) Spearman correlation matrix of the relative abundances of bacterial families and fungal taxa of interest. q-values corrected by false-discovery (FDR) algorithm. (n=1554 samples)

### Impact of intestinal fungal dysbiosis on allo-HCT survival

Finally, we examined whether intestinal fungal dysbiosis in the pre-engraftment period (Day 0 to engraftment) could predict adverse outcomes in allo-HCT patients. We defined fungal dysbiosis as the presence of culturable fecal fungi or the expansion and domination of a single fungal species. Patients with at least one fungal culture-positive fecal sample in the pre-engraftment period had worse overall survival compared to patients without viable fungi in any specimen (Fig 5a). The lower overall survival in patients with viable fungi in fecal specimens was due to a higher transplant-associated mortality (Fig. 5b).

**Figure 5:**
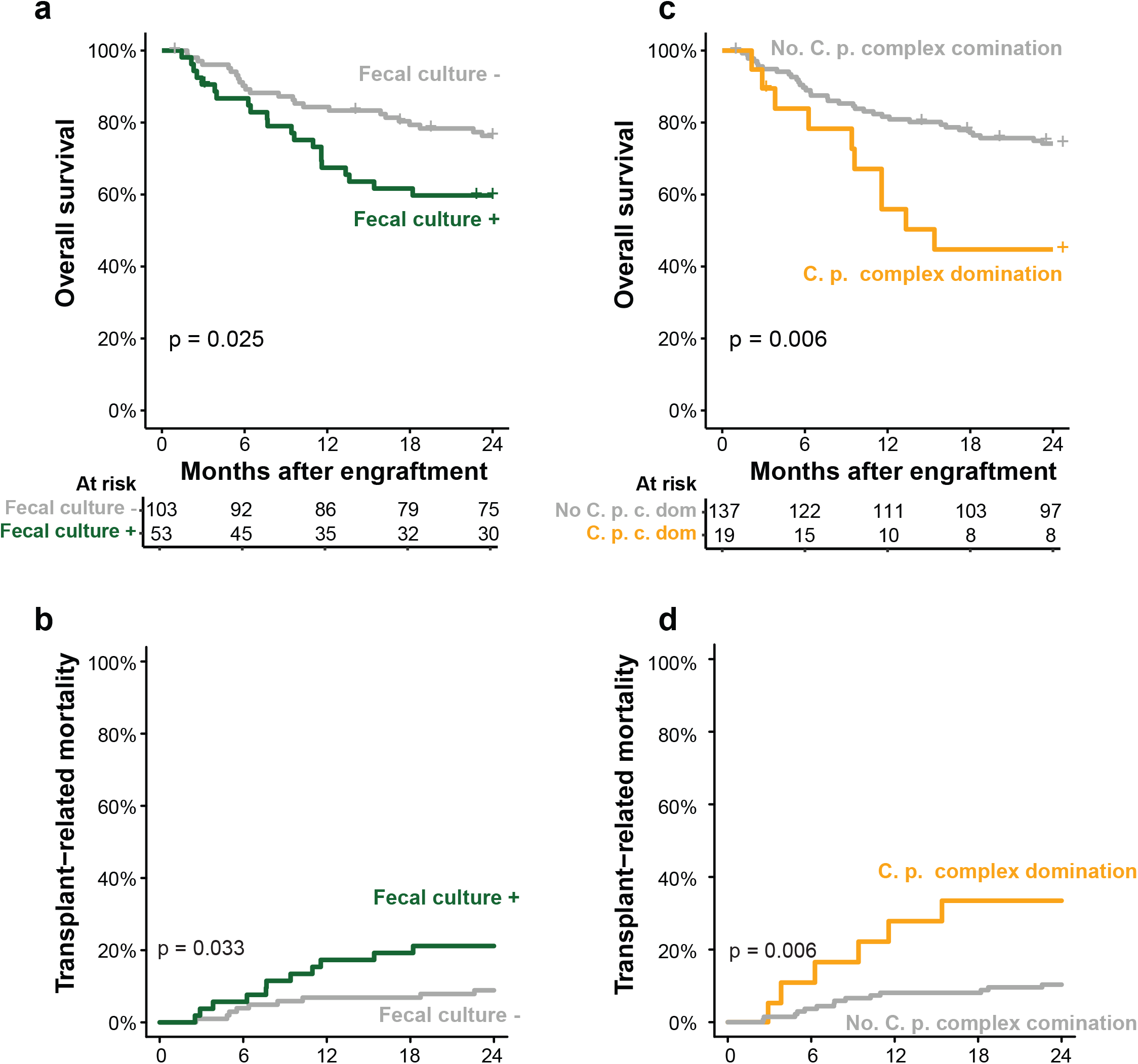
The intestinal fungal mycobiota and clinical outcomes. (a) Overall survival and (b) transplant-related mortality in relation to viable fungi recovered from samples within the pre-engraftment period (days 0 to the first day of an absolute neutrophil count > 500/mm^3^). (c) Overall survival and (d) transplant-related mortality in relation to *C. parapsilosis* complex domination (≥90% relative abundance) within the pre-engraftment period (days 0 to the first day of an absolute neutrophil count > 500/mm^3^). (a, c) Kaplan-Meier curves tested by log-rank test. (b, d) Cumulative incidence curves with relapse and progression of primary disease as competing interest. Differences tested by Gray’s test. See also Extended data Fig. 3.

To assess the impact of a different threshold of relative abundance to define a fungal domination event, we calculated the median relative abundance of the most common taxon in all samples during the pre-engraftment period (82%) and re-examined the clinical outcomes using this alternate threshold. We found that incorporating the alternate threshold in our analyses did not alter the association between fungal dysbiosis and overall survival or between fungal dysbiosis and transplant-related mortality (Extended data Fig. 3a and Extended data Fig. 3b).

Since *C. parapsilosis* complex was most commonly detectable as viable cultures, we tested whether domination with a species belonging to the *C. parapsilosis* complex was similarly predictive of adverse clinical outcomes. Patients with pre-engraftment domination by *C. parapsilosis* complex species had a worse overall survival and higher transplant-related mortality than patients with no pre-engraftment domination by *C. parapsilosis* complex species (Fig. 5C and 5D). In contrast, patients with *C. albicans* domination did not have a lower probability of overall survival or a higher probability of transplant-related mortality compared to patients without a fungal domination event (Extended data Fig. 3c and Extended Fig. 3d).

*C. parapsilosis* complex domination remained associated with worse overall survival even after adjusting for age, comorbidities, graft type, primary disease, and receipt of anti-anaerobic antibiotics, all of which affect outcomes in allo-HCT patients (Table 2). Three *Candida parapsilosis* complex bloodstream infections occurred in the cohort during the study period. All three patients recovered from candidemia. All three episodes were preceded by *Candida parapsilosis* complex intestinal domination (observed in 19/156 patients, 12% incidence; 16% (3/19) incidence of candidemia among patients with *C. parapsilosis* complex domination), as expected from a prior study ^15^.

**Table 2:** Cox proportional hazard model of overall survival.

| Characteristics | Univariate HR<br>(95%CI) | P | Adjusted HR<br>(95%CI) | P |
| --- | --- | --- | --- | --- |
| *C. parapsilosis* complex domination |  |  |  |  |
| - No | 1.00 |  | 1.00 |  |
| - Yes | 2.62 (1.29-5.29) | 0.008 | 2.21 (1.03-4.77) | 0.042 |
| Sex |  |  |  |  |
| - Female | 1.00 |  |  |  |
| - Male | 0.70 (0.39-1.26) | 0.235 |  |  |
| Age (per year increase) | 1.05 (1.01-1.08) | 0.016 | 1.07 (1.03-1.11) | <0.001 |
| HCT-CI (>2 vs 0-2) |  |  |  |  |
| - 0-2 | 1.00 |  | 1.00 |  |
| - >2 | 3.64 (1.88-7.07) | <0.001 | 2.70 (1.37-5.30) | 0.004 |
| Graft type |  |  | - | - |
| - Unmodified <sup>a</sup> | 1.00 |  |  |  |
| - T-cell depleted | 0.85 (0.45-1.62) | 0.433 |  |  |
| Disease |  |  |  |  |
| - Acute Leukemia | 1.00 |  | 1.00 |  |
| - Lymphoma | 0.48 (0.19-1.25) | 0.125 | 0.57 (0.22-1.50) | 0.254 |
| - MDS | 0.84 (0.40-1.77) | 0.545 | 0.63 (0.29-1.34) | 0.203 |
| - other | 0.10 (0.01-0.73) | 0.023 | 0.08 (0.01-0.59) | 0.013 |
| Carbapenem or Piperacillin/Tazobactam |  |  |  |  |
| - No | 1.00 |  | 1.00 |  |
| - Yes | 2.70 (1.33-5.46) | 0.006 | 2.86 (1.34-6.14) | 0.007 |

Patients were classified as having *Candida parapsilosis* complex domination, if they had at least one sample with 90% *Candida parapsilosis* relative abundance or above between days 0 and the day of engraftment. They were classified as having received a carbapenem or piperacillin/tazobactam if they received at least one dose between days 0 and the day of engraftment. <sup>a</sup> Unmodified PBSCs, unmodified BM, and cord grafts were grouped together.

In summary, fungal dysbiosis in allo-HCT patients is characterized by domination of *C. parapsilosis* complex species in conjunction with expansion of culturable intestinal fungi. This fungal expansion and domination were facilitated by a reduction in the intestinal bacterial density. Patients with *C. parapsilosis* and patients with culturable intestinal fungi had a substantially lower overall survival and higher transplant-related mortality after allo-HCT.

## Discussion

In this study, we characterized the dynamics of the fungal intestinal community during allo-HCT by high-density longitudinal sampling. The mycobiota had highly variable densities and compositions across time during HCT. This result is consistent with findings from the Human Microbiome Project, which also demonstrated higher intra- and interindividual variability of intestinal fungi compared to bacteria in healthy individuals ^26,27^. A large proportion of fungal DNA and fungi detected in feces from healthy adults is thought to be due to transient passage rather than direct colonization. Specifically, *C. albicans* in fecal samples has been linked to oral hygiene, while *S. cerevisiae* has been linked to food sources ^27^.

While fungal density did not change over time on a global scale in our cohort, more viable fungi were cultured from samples in the period after HCT. The vast majority of culturable fungi consisted of *C. parapsilosis* complex species, with concurrent expansion observed by ITS1 amplicon sequencing. In contrast, expansion of *C. albicans* ITS1 sequences did not correlate with the presence of culturable fungi. While the efficiency of retrieving culturable fungi may have been reduced due to one cycle of freeze-thawing, sample storage conditions were unlikely to differentially affect *C. parapsilosis* complex and other closely related *Candida* species. The *C. albicans* ITS1 amplicon sequences may have originated from oral or intestinal communities that were impacted by the routine use of micafungin as antifungal prophylaxis during the pre-engraftment allo-HCT period, prior to advancing subjects to azole-based prophylaxis. *C. parapsilosis* complex species are known to have an inherently higher minimal inhibitory concentration against micafungin and other echinocandin drugs compared to other *Candida* species ^28,29^. This phenotype is due to a chromosomal P660A mutation in the *fks1* gene ^30^, though *C. parapsilosis* complex species are not intrinsically resistant to echinocandin drugs. Our findings indicate that *C. parapsilosis* complex species were able to stably colonize the gut in a subset of allo-HCT patients that were treated with micafungin-based antifungal prophylaxis. Antifungal prophylaxis is a standard allo-HCT practice since it was shown over 25 years ago that administration of prophylaxis improves patient survival ^4,5^. Thus, it is not feasible to conduct mycobiota studies during allo-HCT without antifungal prophylaxis. It is possible that different prophylaxis strategies employed across HCT centers worldwide may lead to different *Candida* species domination and fecal culture positivity that in turn may be associated with adverse clinical outcomes.

Colonization by *C. parapsilosis* complex species in the pre-engraftment period was associated with significantly reduced overall survival and higher transplant-related mortality. This association was independent of *Candida* bloodstream infections, although all *Candida* bloodstream infections occurred in the context of intestinal *Candida* expansion, consistent with recent findings ^15^. The presence of culturable *Candida* in oral or fecal samples within the first 10 days after hospitalization of allo-HCT patients has previously been linked to a higher incidence of acute graft-versus-host disease (GvHD, grade II to IV) ^17^. Of note, in that study, subjects did not receive routine antifungal prophylaxis, which had previously been shown to independently reduce the occurrence of severe intestinal GvHD (grades III and IV) ^16^. Since only twelve patients (8%) in our cohort developed acute GvHD of grades III or IV, this study was not powered to detect differences in the incidence of GvHD or GvHD-related mortality.

Recently, profound injury to the intestinal bacterial microbiota during allo-HCT has been linked to worse clinical outcomes in a large multicenter study, including patients from our center^7^. The use of carbapenem drugs and piperacillin-tazobactam diminishes the bacterial intestinal density and modulates the microbiota composition ^8^. Correspondingly, the use of these antibiotics is associated higher transplant-related and higher GvHD-related mortality in allo-HCT patients ^31,32^. In the context of reduced bacterial diversity and under antibiotic selection pressure, bacterial pathobionts are able to expand in the intestine of allo-HCT patients, such as *Enterococcus* and Enterobacteriaceae ^9-14^. Intestinal enterococcal domination has been linked to lower overall survival in allo-HCT patients and been causally involved in the development of acute GvHD ^12^.

Intriguingly, co-domination of enterococci and *C. parapsilosis* was infrequent in our cohort. Prior use of carbapenems or piperacillin-tazobactam, antibiotics that target anaerobic bacteria, only had a small negative confounding effect on the association of *C. parapsilosis* complex domination and overall survival. Thus, the use of broad-spectrum antibiotics is likely to negatively affect patient outcomes by multiple parallel effects, such as loss of bacterial diversity, expansion of bacterial pathobionts, or, as our data suggest, expansion and intestinal colonization by fungal pathobionts.

*C. parapsilosis* complex expansion and an increase in fungal burden occurred in the context of severely diminished bacterial densities and a relative increase of bacterial genera not commonly seen at high abundances in the core intestinal microbiota, such as *Staphylococcus*^33^. This complements our recent findings that patients with *Candida* expansion prior to development of bloodstream infections have a marked loss of intestinal anaerobic bacterial communities ^15^. The findings are also in line with the finding that antibiotic treatment is a prerequisite for stable gut colonization with most *C. albicans* strains in mice ^34^. Metabolites in human feces can inhibit the growth of *C. albicans, C. parapsilosis*, and other fungi in vitro ^35^. The presence of specific anaerobic bacteria in the mouse intestinal microbiota, such as clostridial *Firmicutes* and *Bacteroides thetaiotamicron*, can confer colonization resistance against murine *Candida albicans* colonization ^36,37^. Other intestinal bacteria, such as *Lactobacillus rhamnosus* GG and *E. fecalis* can inhibit *C. albicans* hyphal formation and decrease its virulence ^38,39^. However, since *C. parapsilosis* complex species do not form hyphae, other trans-kingdom interactions may be at play. Determination of the specific mediators of colonization resistance against pathogenic fungi, specifically non-*albicans Candida* species may allow the development of prophylactic probiotic strategies to counter the detrimental effects of antibacterial therapy.

Our data support a model in which the use of broad-spectrum antibiotics and the loss of the bacterial flora promote *C. parapsilosis* complex species expansion under the selective pressure of echinocandin drugs. Fungal overgrowth by these species is in turn associated with worse clinical outcomes in allo-HCT patients. Identifying and targeting fungal dysbiosis in the context of allo-HCT may improve patient outcomes and highlights the clinical importance of the mycobiota in transplantation as therapy for cancer or organ failure.

## Online methods

### Data availability

Fungal ITS 1 and bacterial 16S amplicon sequences will be uploaded to NCBI SRA upon acceptance for publication. Requests for clinical data and materials will be reviewed by Memorial Sloan Kettering to determine if they are subject to intellectual property or confidentiality obligations. Data and materials that can be shared will be released via a material transfer or data-sharing agreements.

### Inclusion Criteria and Sample Collection

Adult patients that undergo allo-HCT at MSKCC are invited to participate in a biospecimen protocol that prospectively and repeatedly collects fecal samples over the course of allo-HCT ^10^. To be included in the current study, patients had to undergo allo-HCT in 2017 or 2018, have a successful cell engraftment, and have at least one fecal sample available in the repository dating between the day of HCT and the day of engraftment. A portion of the patients and samples in this study have been previously reported in studies of the bacterial microbiota ^7,12^. Of this patient pool, we randomly selected 156 patients using the *sample* function in R. All available fecal samples of the selected patients collected between ten days prior to 30 days after HCT were processed. Data on age and sex of included patients can be found in Table 1. All patients provided written informed consent for biospecimen collection. The fecal biospecimen repository and this study protocol were approved by the MSKCC institutional review board. The study was performed in concordance with the Declaration of Helsinki.

### Analysis of fecal samples

After homogenization in 150μl of sterile PBS, 50□μl of liquid stool or 50μg of formed stool were plated on Sabouraud dextrose agar plates complemented with 0.01 mg/ml vancomycin and 0.1 mg/ml gentamicin to culture viable fungi for 48h at 37□°C ^15^. Single colonies were counted to calculate colony-forming units (cfu). Mold colonies were not further classified. Yeast colonies were picked and stored in glycerol stocks at -80°C. Species identification was performed via Sanger sequencing of ITS 1 and comparison of the resulting sequences to the NCBI GenBank nucleotide collection. Our culture-based approach was not optimized to recover viable *Malassezia* species, a major constituent of the skin mycobiota ^40^, due to the requirements for fatty acid supplementation in culture media ^41^. However, *Malassezia* domination by ITS1 amplicon sequencing was observed in only two instances (0.16% of fecal samples), suggesting that very rare events of *Malassezia* intestinal domination did not impact clinical outcomes. For fungal DNA extraction, aliquots of 200-300μg of solid or 200-300μl of liquid stool were weighed. They were then preprocessed with 500□μl of lyticase buffer (50□mM Tris, pH 7.6, 10□mM ethylenediaminetetraacetic acid, 28□mM β-mercaptoethanol) and 100□U lyticase. 0.2ml 0.1-mm silicone beads and 0.3ml 0.5mm beads were added prior to vortexing and incubating the samples for 30 minutes at 32□°C. After adding 30□μl of 4□M NaCl, 47□μl of 2□M Tris-HCl, pH 8.0, 13□μl of 0.5□M ethylenediaminetetraacetic acid, 200□μl of 20% sodium dodecylsulfate and 500□μl of phenol chloroform, the samples were homogenized by bead-beating for 2□min. Detritus was spun down at 13,300g at 4□°C for 5□min. The upper aqueous layer was recovered and added to a mix of 100 μl DEB buffer (0.2M NaCl, 0.2M Tris, pH 8.0, 0.5M EDTA) and 500 μl phenol-chloroform. After inverting and spinning down, the upper aqueous layer was again recovered. This process was repeated for a total of three times. After the final round of phenol-chloroform 200□μl of the aqueous phase was mixed with 200□μl of buffer AL (QIAamp DNA Mini Kit) and 200□μl of 100% ethanol. Further processing was according to the QIAmp DNA Mini Kit instructions. DNA was eluted in 80□μl of ultrapure water.

### Quantification of fungal and bacterial DNA

The amount of fungal and bacterial DNA was quantified via qPCR. 18S copy numbers were determined using the FungiQuant protocol ^22^. The reactions were performed with a TaqMan Fast Advanced Master Mix in a 96-well plate with 10 μl per well and using 1 μl of DNA eluate. Working concentrations were 20μM for the ITS1F and ITS1R primers, and 1μM for the probe. A standard curve was obtained by serial dilutions of genomic DNA from the reference strain C. albicans SC5314. The copy number in the stock was calculated at http://cels.uri.edu/gsc/cndna.html. After 2 minutes at 50□°C and 10 minutes at 95 °C, cycling conditions were15 seconds at 95□°C and 1 minute at 60□°C for a total of 40 cycles. 16S copy number were determined using a protocol based on the DyNAmo SYBR Green rtPCR Kit (Finnzymes) and the broad-range bacterial 16S primers 563F and 926Rb at 0.2mM. Cycling conditions were 10 minutes at 95□°C followed by 40 cycles of 95□°C for 30 s, 52□°C for 30 s and 72□°C for 1 min. Standard curves were obtained by serial dilution of the PCR blunt vector (Invitrogen) containing one copy of the 16S rRNA gene derived from a member of the Porphyromonadaceae family. For both 18S and 16S copy numbers, values were normalized to the fecal weight input.

### Amplicon preparation, sequencing, and data processing

ITS1 and 16S (V4-V5) amplicon preparation were performed as reported previously by our group ^15,42^. For both the 16S and ITS1-specific forward and reverse primers were modified with unique 12-base Golay barcodes preceding the primer for parallel sequencing of up to 60 samples per sequencing pool and subsequent sample identification. To offset primer sequencing, additional 1 to 8 nucleotides were placed in front of the barcode ^42^.

For ITS 1 amplicon preparation 1 μl of purified sample eluate template DNA (at 1:1, 1:5, and 1:25 dilutions) was amplified with the ITS1F and ITS1R primers (1.25μM) and Phusion enzyme for 35 cycles: 30 s at 98°C, 30 s at 53°C, and 30s at 72°C. The resulting three PCR amplicon mixes per sample were pooled and purified using the Axygen AxyPrep MagKit Mag PCR Clean-Up Kit according to the manufacturer’s protocol. For 16S amplicon preparation each sample, duplicate 50-μl PCRs were performed. 50 ng of purified DNA were amplified with 2.5 U Platinum Taq DNA polymerase, and 0.5 mM of primers 563F and 926R at 94°C for 3 min, followed by 27 cycles of 94°C for 50 s, 51°C for 30 s, and 72°C for 1 min and a final elongation step at 72°C for 5 min.

Amplicons were purified using the QIAquick PCR Purification Kit (Qiagen) after pooling of replicates. DNA content in the 16S and ITS amplicon mixes was measured by Qubit Assay. Equimolar amounts of DNA were pooled and sent to the MSKCC Integrated Genomics Operation for library preparation and sequencing on the Illumina MiSeq platform using PE300 for ITS1 and PE250 for 16S setting.

### ASV Construction and taxonomic identification

Raw reads were demultiplexed on a per-sample basis according to the unique Golay barcodes. After primer and barcode removal, the DADA2 pipeline was used for quality filtering and inference of exact amplicon sequencing variants (ASVs). For 16S amplicons, the default values in DADA2 were used, for ITS1 amplicons, we used a truncation length of 257 and a maximum expected (*maxEE*) error of 5. To accommodate ITS1 segments exceeding PE300 settings, we concatenated forward and reverse reads only in the absence of an overlap.

Taxonomy was assigned at species level with an algorithm based on BLAST with NCBI RefSeq RNA for 16S and UNITE database for ITS1 as training sets, respectively^43,44^. ASV tables, taxonomy, and sample metadata were compiled in a phyloseq object in R ^45^. Low-quality samples with less than 200 reads were removed from further analysis, both for ITS 1 and 16S.

### Statistical Analyses

All analyses were performed in R. The statistical methods used are described in the figure legends. The limits of the boxplots in the figures correspond to the lower and upper quartiles, with the line in-between representing the median. The whiskers correspond to the range excluding outliers. Hierarchical clustering of samples was performed by using weighted unifrac distances at the genus level (as determined by the taxonomy in the UNITE database). Fungal and bacterial diversity were estimated by the Inverse Simpson Index. Stability between sample pairs of the same patient was estimated by calculating the Chao-Jaccard index between these two samples. For all survival and clinical outcome analyses, the exposure of interest had to occur between the day of cell infusion and the first day the absolute neutrophil count returned over 0.5 (engraftment). Outcome analyses started at engraftment. As only patients that had engrafted were included in the study, no patient died before engraftment. To compare overall survival Kaplan-Meier curves were plotted and between-group differences tested by log-rank test. Transplant-related mortality was defined as any death except deaths due to relapses or progressive primary disease. To assess transplant-related mortality, cumulative incidences were plotted with relapse and progressive disease as competing interests using the *cmprsk* package in R. Between group differences were tested by Gray test. For both, overall survival and transplant-related mortality, patients were censored at 2 years after engraftment or at last contact, whichever was shorter. To estimate the hazard ratio of overall survival in patients with and without C. parapsilosis domination, a Cox proportional hazard model was used. Covariates tested in the univariate analysis included age at transplantation, sex, graft type, primary disease, HCT-CI, and receipt of piperacillin-tazobactam or a carbapenem in the pre-engraftment period. Covariates associated with overall survival at p=0.2 were included in a multivariate model. Piperacillin-tazobactam and carbapenems were chosen as they have been previously shown to have the largest detrimental impact on the anaerobic intestinal microbiota and has been associated with worse clinical outcomes in allo-HCT patients ^8,32^.

To correlate relative bacterial and fungal abundances, we grouped bacteria at the genus level and looked at three fungal taxonomic groups: *C. parapsilosis* complex, other species of the Candida genus, and S. cerevisiae. Cross-correlation was performed with the *associate* function in the R package *microbiome* by using Spearman correlation and adjusting p-values by the false discovery rate (FDR) algorithm.

## Data Availability

All sequence data have been submitted to SRA and will be released upon final acceptance of the manuscript. Requests for clinical data and materials will be reviewed by Memorial Sloan Kettering to determine if they are subject to intellectual property or confidentiality obligations. Data and materials that can be shared will be released via a material transfer or data-sharing agreements.

## Acknowledgments

This work was supported by Deutsche Forschungsgemeinschaft (DFG, German Research Foundation) grant RO-5328/2 (T.R.), National Institutes of Health (NIH) grants R01 AI093808 (T.M.H.), R21 AI105617 (T.M.H.), R21 AI156157 (T.M.H.), U01 AI124275 (J.B.X.), R01 AI137269 (J.B.X. and Y.T.), K08 HL143189 (J.U.P.), R01 CA228358 (M.R.M.v.d.B.), R01 CA228308 (M.R.M.v.d.B.), P01 CA023766 (M.R.M.v.d.B.), R01 HL125571 (M.R.M.v.d.B.), R01 HL123340 (M.R.M.v.d.B.), P01 AG052359 (Project 2, M.R.M.v.d.B.), a Takeda Science Foundation-Fellowship (K.Y.M.), an Investigator in the Pathogenesis of Infectious Diseases Award from the Burroughs Wellcome Fund (T.M.H.), the Ludwig Center for Cancer Immunotherapy (T.M.H.), a Tri-Institutional Stem Cell Initiative award 2016-013 (M.R.M.v.d.B.), the Lymphoma Foundation (M.R.M.v.d.B.), the Parker Institute for Cancer Immunotherapy at MSKCC (M.R.M.v.d.B., K.A.M., and J.U.P.), the Susan and Peter Solomon Divisional Genomics Program (T.M.H. and M.R.M.v.d.B.), and the DKMS (K.A.M.). All authors were supported by NIH P30 CA008748 (Cancer Center Core Grant).

## Author contributions

Study conception: T.R., B.Z. & T.M.H., Clinical data collection: D.M.P., M.A.P, M.R.M.vdB., K.A.M., J.U.P, Y.T., T.M.H, Data analysis and visualization: T.R., with assistance from B.Z., J.B.X., Y.T., J.U.P., Sample processing: T.R.,B.Z.,M.G.,N.T.,K.Y.M.,E.F.,L.A.A., R.J.W.

Manuscript writing: T.R. & T.M.H. All co-authors reviewed and edited the manuscript.

## Figure Legends

**Extended data figure 1: Impact of culture-positivity and TPN administration on fungal and bacterial microbiota dynamics during allo-HCT, related to Fig. 1**.

(a) Comparison of fungal density (n = 246 pre-transplant samples and n = 978 post-transplant samples) and (b) fungal diversity (n = 251 pre-transplant samples and n = 1028 post-transplant samples) between the pre- (days -10 to -1) and the post-transplant period (days 0 to 30). (c) Comparison of bacterial density (n = 204 pre-transplant samples and n = 749 post-transplant samples) and (d) diversity (n = 286 pre-transplant samples and n = 939 post-transplant samples) between the pre- and post-transplant period. Statistical tests (a-d): P-values issued from generalized estimating equations with patient ID as cluster variable are shown.

**Extended data Fig. 2: Taxonomic composition and relation to culture-positivity, related to Fig. 3:** (a) Taxonomic composition of culture-positive samples by ITS (top ribbon) and species cultured from the respective samples (bottom ribbon). (b) Comparison of fungal similarity (1-Chao-Jaccard index) according to presence of fungal domination status of the earliest sample. Each dot represents a sample pair of an individual patient collected one day apart (n= 516 sample pairs). (c) Comparison of fungal similarity (1-Chao-Jaccard index) according to the taxa responsible for fungal domination in the earliest samples. Each dot represents a sample pair of an individual patient collected one day apart (n= 88 sample pairs). (b-c) P-values issued from generalized estimating equations with patient ID as cluster variable are shown.

**Extended data Fig. 3: Supplementary data for clinical outcomes and the fungal intestinal microbiota, related to Fig. 5**.

(a) Overall survival and (b) transplant-related mortality in relation to a relative abundance of *C. parapsilosis* complex ≥ 82% within the pre-engraftment period (days 0 to the first day of an absolute neutrophil count > 500/mm^3^). (a) Kaplan-Meier curves tested by log-rank test. (b) Cumulative incidence curves with relapse and progression of primary disease as competing interest. Differences tested by Gray’s test. (c) Overall survival and (d) transplant-related mortality in relation to domination by *C. albicans* within the pre-engraftment period (days 0 to the first day of an absolute neutrophil count > 500/mm^3^). (c) Kaplan-Meier curves tested by log-rank test. (d) Cumulative incidence curves with relapse and progression of primary disease as competing interest. Differences tested by Gray’s test.

