## Supplementary figures and images for "Intestinal fungal dynamics and linkage to hematopoietic cell transplantation outcomes"

### Figure S1

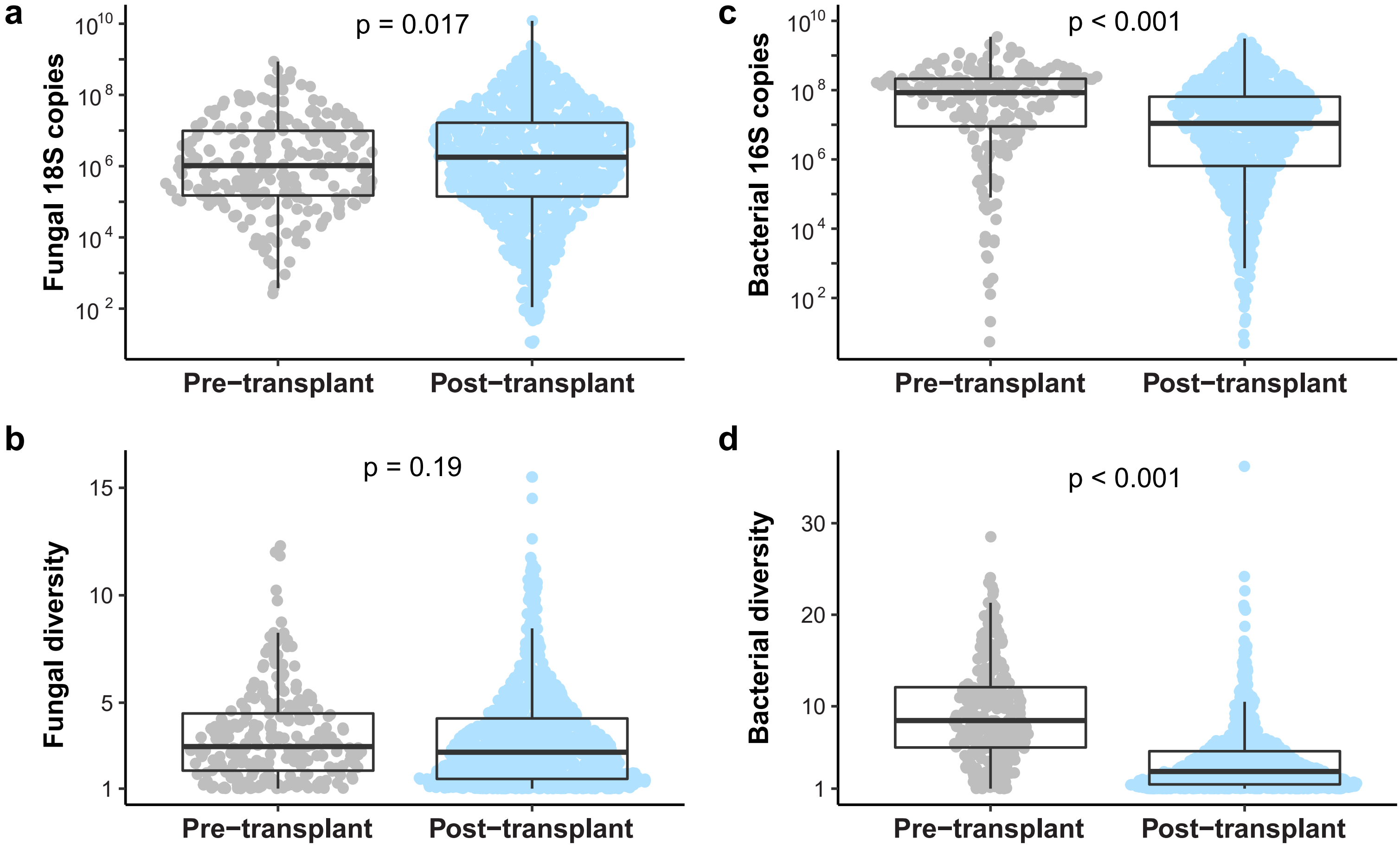


**Ext. data Fig.S1**

### Figure S2

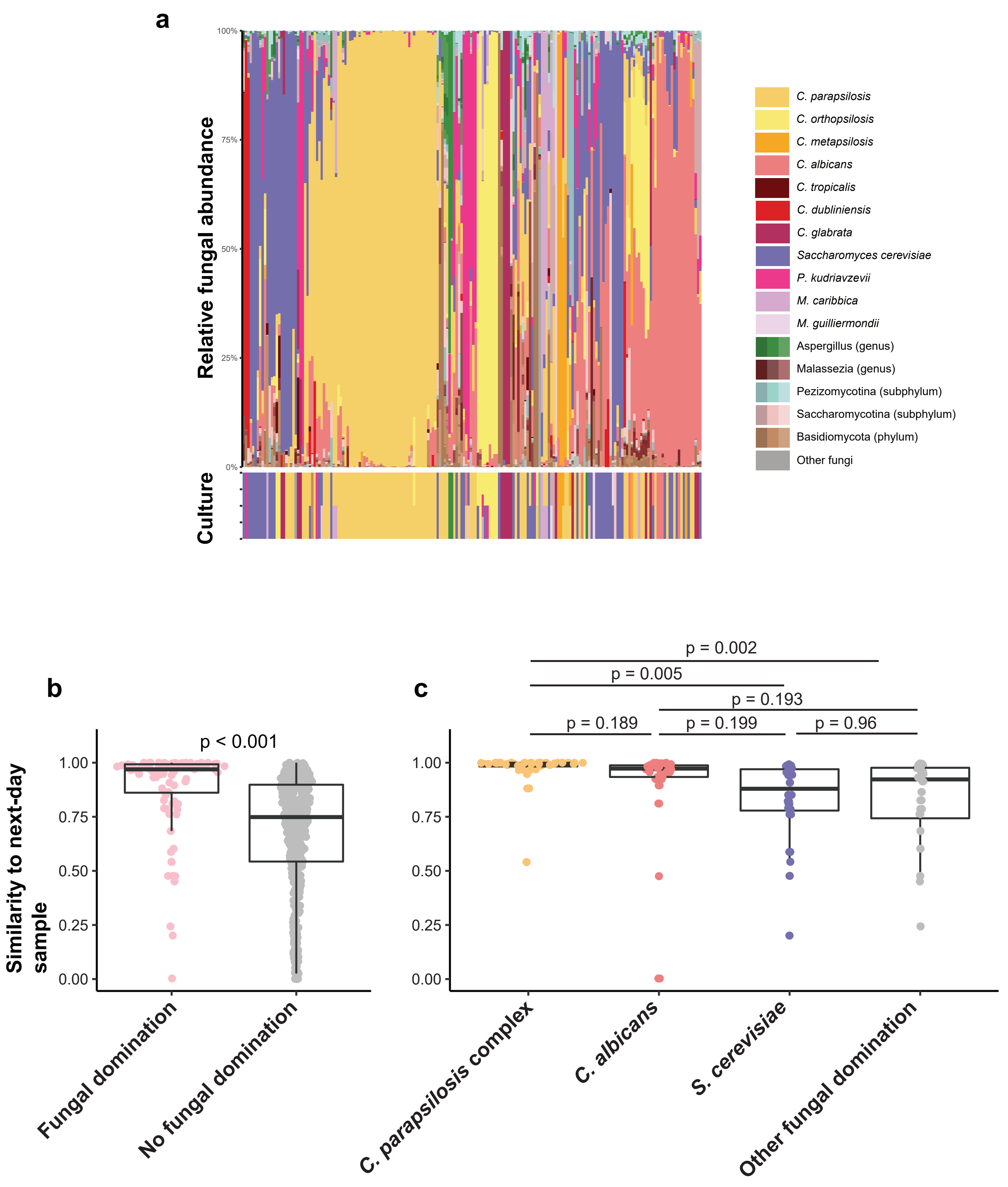


**Ext. data Fig. 2**

### Figure S3

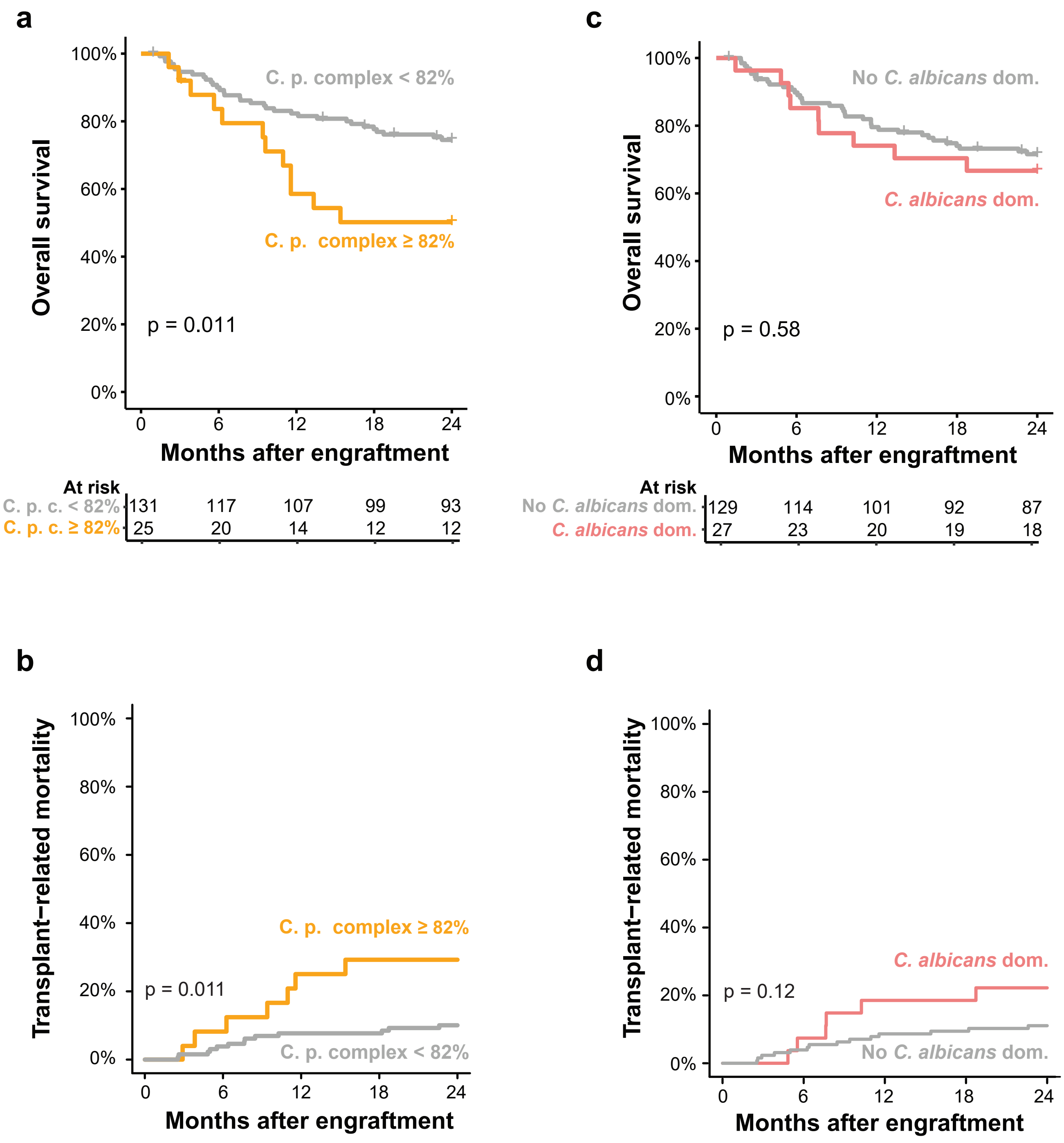


**Ext. data Fig. 3**
